# A cohort study of 223 patients explores the clinical risk factors for the severity diagnosis of COVID-19

**DOI:** 10.1101/2020.04.18.20070656

**Authors:** Yongsheng Huang, Xiaoyu Lyu, Dan Li, Yujun Wang, Lin Wang, Wenbin Zou, Yingxin Wei, Xiaowei Wu

## Abstract

**BACKGROUND:** Coronavirus Disease 2019 (COVID-19) has recently become a public emergency and a worldwide pandemic. The clinical symptoms of severe and non-severe patients vary, and the case-fatality rate (CFR) in severe COVID-19 patients is very high. However, the information on the risk factors associated with the severity of COVID-19 and of their prognostic potential is limited.

**METHODS:** In this retrospective study, the clinical characteristics, laboratory findings, treatment and outcome data were collected and analyzed from 223 COVID-19 patients stratified into 125 non-severe patients and 98 severe patients. In addition, a pooled large-scale meta-analysis of 1646 cases was performed.

**RESULTS:** We found that the age, gender and comorbidities are the common risk factors associated with the severity of COVID-19. For the diagnosis markers, we found that the levels of D-dimer, C-reactive protein (CRP), lactate dehydrogenase (LDH), procalcitonin (PCT) were significantly higher in severe group compared with the non-severe group on admission (D-Dimer: 87.3% vs. 35.3%, *P*<0.001; CRP, 65.1% vs. 13.5%, *P*<0.001; LDH: 83.9% vs. 22.2%, *P*<0.001; PCT: 35.1% vs. 2.2%, *P*<0.001), while the levels of aspartate aminotransferase (ASP) and creatinine kinase (CK) were only mildly increased. We also made a large scale meta-analysis of 1646 cases combined with 4 related literatures, and further confirmed the relationship between the COVID-19 severity and these risk factors. Moreover, we tracked dynamic changes during the process of COVID-19, and found CRP, D-dimer, LDH, PCT kept in high levels in severe patient. Among all these markers, D-dimer increased remarkably in severe patients and mostly related with the case-fatality rate (CFR). We found adjuvant antithrombotic treatment in some severe patients achieved good therapeutic effect in the cohort.

**CONCLUSIONS:** The diagnosis markers CRP, D-dimer, LDH and PCT are associated with severity of COVID-19. Among these markers, D-dimer is sensitive for both severity and CFR of COVID-19. Treatment with heparin or other anticoagulants may be beneficial for COVID-19 patients.

**Funding:** This study was supported by funding from the National Key Research and Development Program of China (2016YFC1302203); Beijing Nova Program (grant number: xx2018040).

**Role of the funding source:** The funding listed above supports this study, but had no role in the design and conduct of the study.

## Introduction

In December 2019, a novel coronavirus disease (COVID-19) caused by Severe Acute Respiratory Syndrome Coronavirus 2 (SARS-CoV-2) infection broke out in Wuhan and spread rapidly throughout China. As of April 18th, 2020, World Health Organization (WHO) reported 2,160,207 COVID-19 cases have been confirmed in most countries or areas, and 146,088 people died from COVID-19(1). The overall COVID-19 case-fatality rate (CFR) has approached about 4%. Human to human transmission has accounted for most if not all of the COVID-19 infections, including the medical personnel in hospital (2, 3). The clinical manifestations of COVID-19 included fever, cough, diarrhea, dyspnea, fatigue and pneumonia (4-6). Most patients with COVID-19 were considered as non-severe patients and the symptoms generally mild. However, the symptoms in about 10% of COVID-19 patients are severe and some progress rapidly to critical conditions, including organ dysfunction, such as acute respiratory distress syndrome (ARDS), acute cardiac injury, acute kidney injury and even death(7, 8). So far, there is no specific medicine or effective vaccines for COVID-19, and the diagnosis methods for the severity of COVID-19 patients were limited. Thus, our objective of this cohort is to explore the risk factors which related with the severity of COVID-19 and put it into practice. In addition, we used meta-analysis to verify these risk factors. Our study would be helpful for setting up different treatment and personal therapy route for COVID-19 patients.

## Results

### Clinical characteristics for the cohort on admission

The general clinical characteristics of 223 COVID-19 cases in our study are shown in **Table 1**. Patients were categorized into 125 non-severe and 98 severe subgroups on admission respectively. In our cohort, the median age in the non-severe population was 52.0 years, interquartile range (IQR) was 26.0 to 65.5, in severe population was 66 years (IQR: 56.3 to 71.0). The age differed significantly between the two subgroups, and 44.8% of severe patients were aged above 60 years. Men are more likely to develop severe COVID-19 than women (66.3% vs. 33.7%). Fever (78.9%), cough (67.7%) and fatigue (56.5) were the most common symptoms, whereas diarrhea (6.7%) was rare. Notably, fever occurred more frequently in the patients in the severe group than the non-severe group (85.7% vs.73.6%). We also found the comorbidities were risk factors associated withe the severity of COVID-19 in our cohort. More severe patients had common chronic diseases than non-severe patients, such as hypertension (38.8% vs. 9.6%) and diabetes (12.2% vs. 5.6%).

**Table 1.**
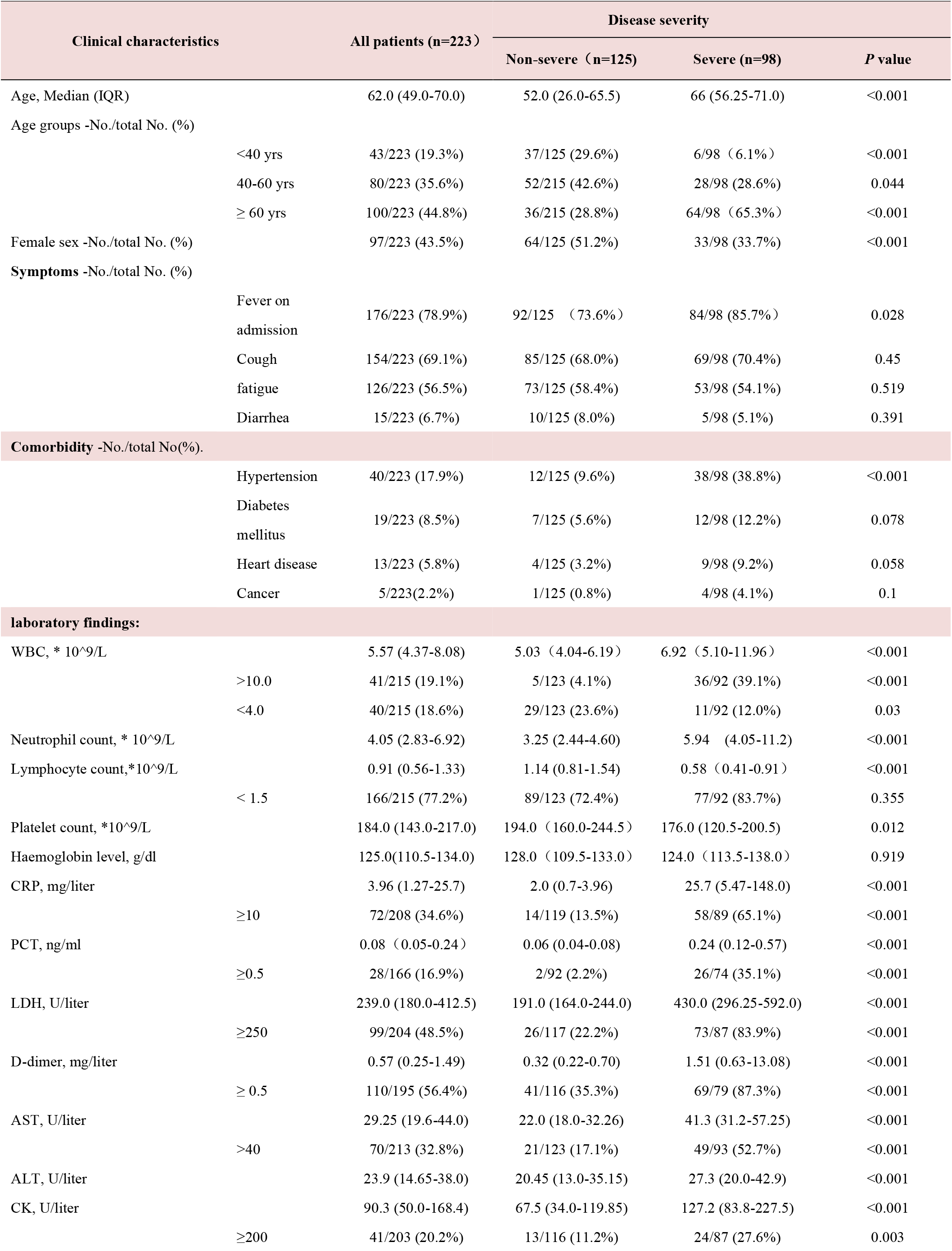
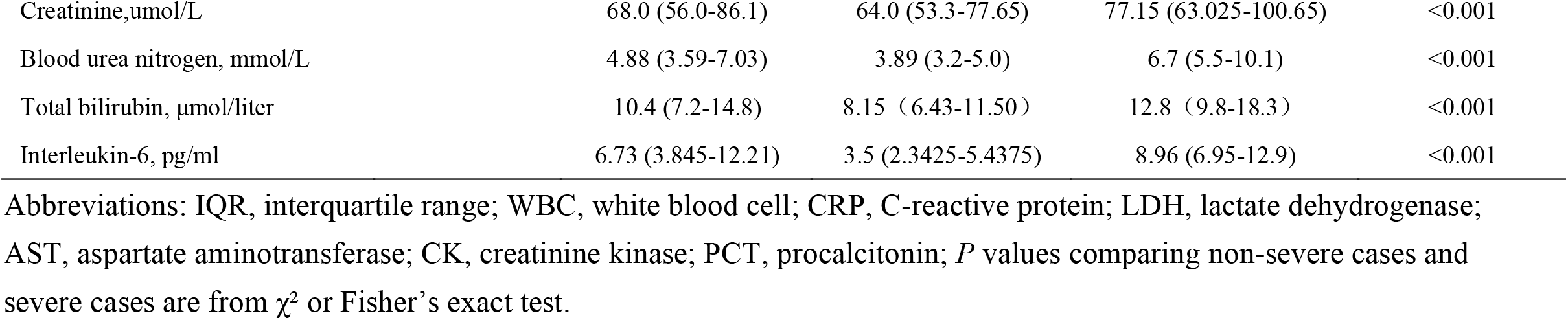
Clinical characteristics and pertinent Laboratory findings of the patients with COVID-19 in the cohort.

| Clinical characteristics | All patients (n=223) | Disease severity |  |  |
| --- | --- | --- | --- | --- |
|  |  | Non-severe (n=125) | Severe (n=98) | P value |
| Age, Median (IQR) | 62.0 (49.0-70.0) | 52.0 (26.0-65.5) | 66 (56.25-71.0) | <0.001 |
| Age groups -No./total No. (%) |  |  |  |  |
| <40 yrs | 43/223 (19.3%) | 37/125 (29.6%) | 6/98 (6.1%) | <0.001 |
| 40-60 yrs | 80/223 (35.6%) | 52/125 (42.6%) | 28/98 (28.6%) | 0.044 |
| ≥ 60 yrs | 100/223 (44.8%) | 36/125 (28.8%) | 64/98 (65.3%) | <0.001 |
| Female sex -No./total No. (%) | 97/223 (43.5%) | 64/125 (51.2%) | 33/98 (33.7%) | <0.001 |
| Symptoms -No./total No. (%) |  |  |  |  |
| Fever on admission | 176/223 (78.9%) | 92/125 (73.6%) | 84/98 (85.7%) | 0.028 |
| Cough | 154/223 (69.1%) | 85/125 (68.0%) | 69/98 (70.4%) | 0.45 |
| fatigue | 126/223 (56.5%) | 73/125 (58.4%) | 53/98 (54.1%) | 0.519 |
| Diarrhea | 15/223 (6.7%) | 10/125 (8.0%) | 5/98 (5.1%) | 0.391 |
| Comorbidity -No./total No(%). |  |  |  |  |
| Hypertension | 40/223 (17.9%) | 12/125 (9.6%) | 38/98 (38.8%) | <0.001 |
| Diabetes mellitus | 19/223 (8.5%) | 7/125 (5.6%) | 12/98 (12.2%) | 0.078 |
| Heart disease | 13/223 (5.8%) | 4/125 (3.2%) | 9/98 (9.2%) | 0.058 |
| Cancer | 5/223(2.2%) | 1/125 (0.8%) | 4/98 (4.1%) | 0.1 |
| laboratory findings: |  |  |  |  |
| WBC, * 10<sup>9</sup>/L | 5.57 (4.37-8.08) | 5.03 (4.04-6.19) | 6.92 (5.10-11.96) | <0.001 |
| >10.0 | 41/215 (19.1%) | 5/123 (4.1%) | 36/92 (39.1%) | <0.001 |
| <4.0 | 40/215 (18.6%) | 29/123 (23.6%) | 11/92 (12.0%) | 0.03 |
| Neutrophil count, * 10<sup>9</sup>/L | 4.05 (2.83-6.92) | 3.25 (2.44-4.60) | 5.94 (4.05-11.2) | <0.001 |
| Lymphocyte count,*10<sup>9</sup>/L | 0.91 (0.56-1.33) | 1.14 (0.81-1.54) | 0.58 (0.41-0.91) | <0.001 |
| < 1.5 | 166/215 (77.2%) | 89/123 (72.4%) | 77/92 (83.7%) | 0.355 |
| Platelet count, *10<sup>9</sup>/L | 184.0 (143.0-217.0) | 194.0 (160.0-244.5) | 176.0 (120.5-200.5) | 0.012 |
| Haemoglobin level, g/dl | 125.0(110.5-134.0) | 128.0 (109.5-133.0) | 124.0 (113.5-138.0) | 0.919 |
| CRP, mg/liter | 3.96 (1.27-25.7) | 2.0 (0.7-3.96) | 25.7 (5.47-148.0) | <0.001 |
| ≥10 | 72/208 (34.6%) | 14/119 (13.5%) | 58/89 (65.1%) | <0.001 |
| PCT, ng/ml | 0.08 (0.05-0.24) | 0.06 (0.04-0.08) | 0.24 (0.12-0.57) | <0.001 |
| ≥0.5 | 28/166 (16.9%) | 2/92 (2.2%) | 26/74 (35.1%) | <0.001 |
| LDH, U/liter | 239.0 (180.0-412.5) | 191.0 (164.0-244.0) | 430.0 (296.25-592.0) | <0.001 |
| ≥250 | 99/204 (48.5%) | 26/117 (22.2%) | 73/87 (83.9%) | <0.001 |
| D-dimer, mg/liter | 0.57 (0.25-1.49) | 0.32 (0.22-0.70) | 1.51 (0.63-13.08) | <0.001 |
| ≥ 0.5 | 110/195 (56.4%) | 41/116 (35.3%) | 69/79 (87.3%) | <0.001 |
| AST, U/liter | 29.25 (19.6-44.0) | 22.0 (18.0-32.26) | 41.3 (31.2-57.25) | <0.001 |
| >40 | 70/213 (32.8%) | 21/123 (17.1%) | 49/93 (52.7%) | <0.001 |
| ALT, U/liter | 23.9 (14.65-38.0) | 20.45 (13.0-35.15) | 27.3 (20.0-42.9) | <0.001 |
| CK, U/liter | 90.3 (50.0-168.4) | 67.5 (34.0-119.85) | 127.2 (83.8-227.5) | <0.001 |
| ≥200 | 41/203 (20.2%) | 13/116 (11.2%) | 24/87 (27.6%) | 0.003 |
| Creatinine,umol/L | 68.0 (56.0-86.1) | 64.0 (53.3-77.65) | 77.15 (63.025-100.65) | <0.001 |
| Blood urea nitrogen, mmol/L | 4.88 (3.59-7.03) | 3.89 (3.2-5.0) | 6.7 (5.5-10.1) | <0.001 |
| Total bilirubin, μmol/liter | 10.4 (7.2-14.8) | 8.15 (6.43-11.50) | 12.8 (9.8-18.3) | <0.001 |
| Interleukin-6, pg/ml | 6.73 (3.845-12.21) | 3.5 (2.3425-5.4375) | 8.96 (6.95-12.9) | <0.001 |

Abbreviations: IQR, interquartile range; WBC, white blood cell; CRP, C-reactive protein; LDH, lactate dehydrogenase; AST, aspartate aminotransferase; CK, creatinine kinase; PCT, procalcitonin; *P* values comparing non-severe cases and severe cases are from $\chi^2$ or Fisher's exact test.

The representative radiologic findings of one patient with non-severe and another three patients with severe COVID-19 were demonstrated in **Figure 1**. Severe cases yielded more prominent radiologic abnormalities on CT than non-severe cases. The laboratory findings on admission were shown in **Table 1**. Of 223 patients, 77.2% of patients had lymphopenia and the severe group is more likely to develop lymphopenia (83.7 vs. 73.4). On admission, severe cases also had other prominent laboratory abnormalities as compared to the non-severe cases (**Table 1**), such as neutrophilia, thrombocytopenia, elevated levels of C-reactive protein (CRP, 65.1% vs. 13.3%, *P*<0.001), D-dimer (D-Dimer: 87.3% vs. 35.3%, *P*<0.001), procalcitonin (PCT: 35.1% vs. 2.2%, *P*<0.001), lactate dehydrogenase (LDH: 83.9% vs. 22.2%, *P*<0.001), aspartate aminotransferase (AST: 52.7 % vs. 11.7%, *P*<0.001) and creatine kinase (CK: 27.6 % vs. 11.2%, *P=*0.003).

**Figure 1:**
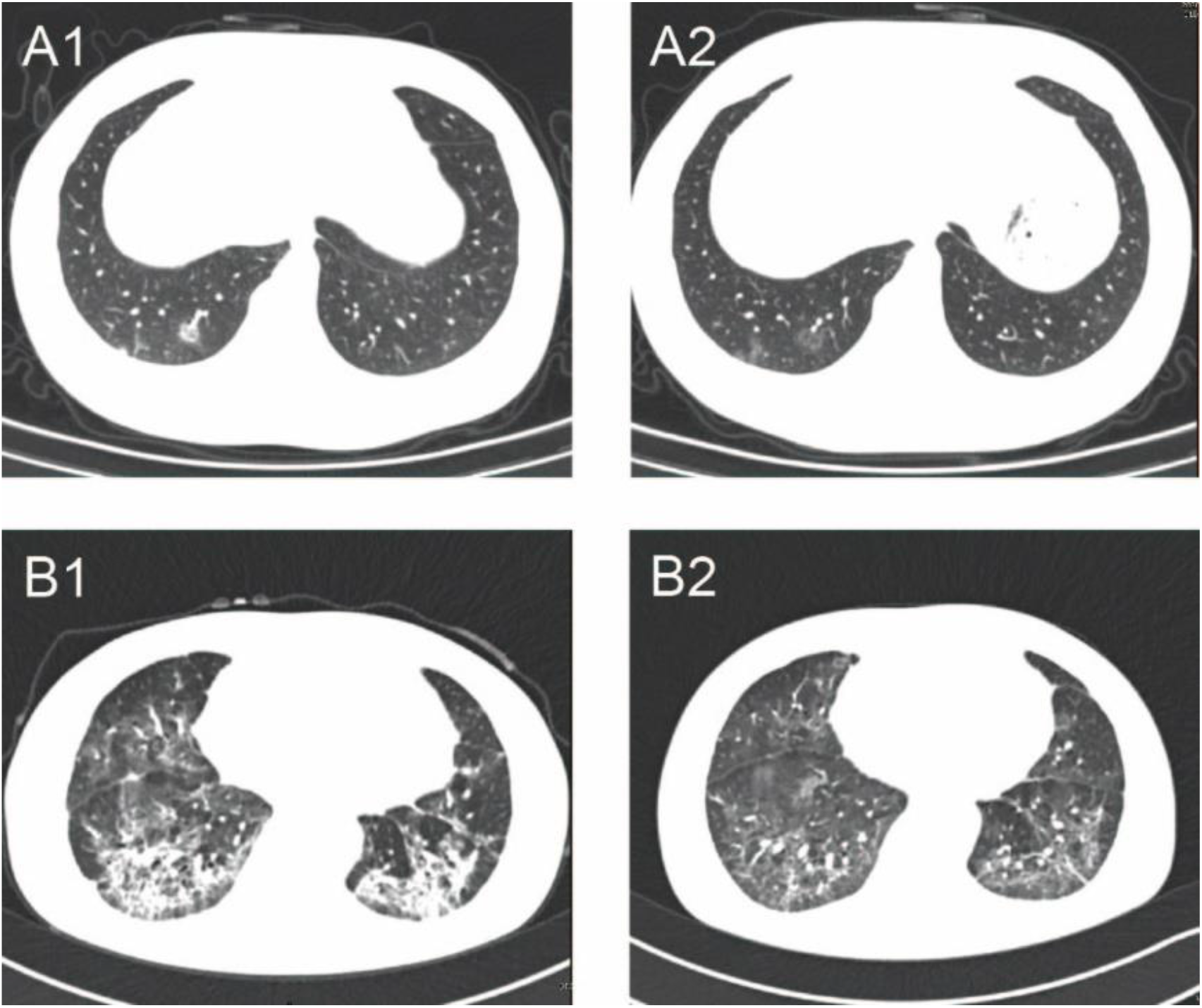
Representative images of the thoracic CT scans showing the changes in COVID-19 patients during the treatment. A. CT images of a non-severe patient. After 5 days of treatment, the foci of infection were observed to be significantly absorbed in a 39-year-old female patient. A1 and A2 were CT images before and after treatment. B. CT images of a severe patient. After three weeks of treatment, significant improvement was observed in large area flaps infection of double lower lung of a 65-year-old female patient. B1 and B2 were CT images before and after treatment, respectively.

### The characteristics of risk factors in the meta-analysis

**Table 2** is the summary of case studies (4, 5, 9, 10) examining the association between clinical characters and COVID-19 in the meta-analysis, dividing into two subtypes: severe and non-severe patients. For each study, we investigated the association between the risk factors and COVID-19, assuming different calculation models. Overall, when all the eligible studies were pooled into the meta-analysis, significant associations were found for elevated levels of CRP [odd ratios (OR) =5.78, 95% Confidence interval (CI): 2.86-11.68, *P*<0.001], D-dimer (OR=4.81, 95%CI: 0.76-30.33, *P=*0.094), PCT (OR=6.51 95%CI:3.48-12.18, *P*<0.001), LDH (OR=4.69, 95%CI: 1.47-14.95, *P*<0.009) and AST (OR=1.98, 95%CI: 1.46-2.70, *P*<0.001) with the severity of COVID-19 (**Table 2, Figure 2**).

**Table 2.** Descriptive clinical risk factors of COVID-19 cases by meta-analysis.

| Studies<br>Risk factors | Guan W. |  | Huang C. |  | Zhang J. |  | Xiao K. |  | Our Study |  | OR(95%CI) | P value |
| --- | --- | --- | --- | --- | --- | --- | --- | --- | --- | --- | --- | --- |
|  | non-severe | severe | non-severe | severe | non-severe | severe | non-severe | severe | non-severe | severe |  |  |
| WBC ↑ | 39 /811 | 19 /167 | 5/27 | 7/13 | 4 /82 | 13 /56 | 4 /107 | 1 /36 | 5 /123 | 36 /92 | 4.59 (3.09-6.80) | <0.001 |
| WBC ↓ | 228 /811 | 102/167 | 9/27 | 1/13 | 18 /82 | 9 /56 | 26 /107 | 11 /36 | 29 /123 | 11 /92 | 0.91 (3.09-6.81) | 0.873 |
| Lymphocyte↓ | 584 /726 | 147 /153 | 15/28 | 11/13 | 58 /82 | 46 /56 | 46 /107 | 20 /36 | 89 /123 | 77 /92 | 5.78 (2.86-11.68) | <0.001 |
| CRP ↑ | 371 /658 | 110 /135 | N/A |  | 12 /43 | 23 /38 | 43 /107 | 29 /36 | 14 /119 | 58 /89 | 5.78 (2.86-11.68) | <0.001 |
| LDH ↑ | 205 /551 | 72 /124 | 17/27 | 12/13 | N/A |  | 28 /107 | 15 /36 | 26 /117 | 73 /87 | 4.69 (1.47-14.95) | 0.009 |
| AST ↑ | 112 /615 | 56 /142 | 1/4 | 8/13 | N/A |  | N/A |  | 21 /123 | 49 /93 | 1.98 (1.46-2.70) | <0.001 |
| CK ↑ | 67 /536 | 23 /121 | 7/27 | 6/13 | 1 /35 | 3 /25 | N/A |  | 13/116 | 24/87 | 2.13 (1.44-3.16) | <0.001 |
| D-dimer ↑ | 195 /451 | 65 /109 | N/A |  | N/A |  | N/A |  | 41 /116 | 69 /79 | 4.81 (0.76-30.33) | <0.001 |
| PCT ↑ | N/A |  | N/A |  | 16 /68 | 25 /50 | 1 /107 | 3 /36 | 2 /92 | 26 /74 | 6.51 (3.48-12.18) | <0.001 |

Abbreviations: WBC, white blood cell; CRP, C-reactive protein; LDH, lactate dehydrogenase; AST, aspartate aminotransferase; CK, creatinine kinase; PCT, procalcitonin; N/A, not applicable. Z test was performed to determine the significance of the pooled OR ( $P \leq 0.05$ suggests a significant OR).

**Figure 2:**
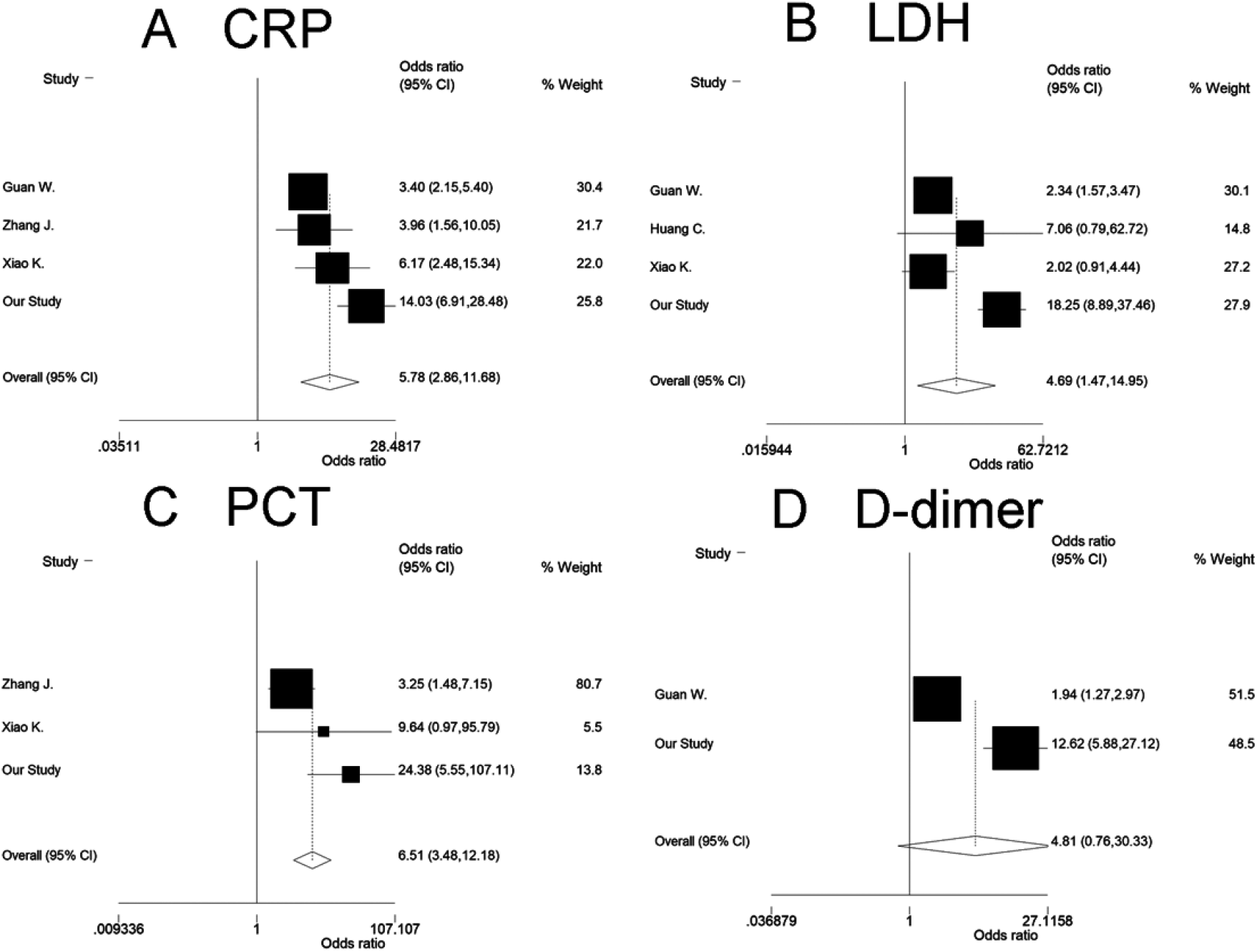
Forest plot of pooled meta-analysis to detect the association between risk factors and severity of COVID-19. Figure shows the association between elevated risk factors and severity of COVID-19 in the meta-analysis: CRP (A), LDH (B), PCT(C) and D-dimer (D). The squares and horizontal lines correspond to the study-specific odd ratios (OR) and 95% confident incidence (CI). The area of the squares reflects the weight. The diamond represents the summary OR and 95% CI. Abbreviations: CRP, C-reactive protein; LDH, lactate dehydrogenase; PCT, procalcitonin.

### Dynamics of risk factors predict progressive severity of COVID-19

We also evaluated the dynamics of clinical risk factors during hospital stay between non-severe and severe COVID-19 patients. Of the risk factors evaluated, we found the levels of D-dimer, CRP, LDH, and PCT are significantly higher in severe patients than in non-severe patients **(Figure3**). Moreover, we found that, while levels of CRP, LDH, and PCT were consistently high in severe patient, D-dimer level still increased in some severe patients, which means it was most likely related to severity **(Figure3**). Strikingly, when further stratified the severe patient into two subgroups: Recovered (R-severe, 31 cases) and Death (D-severe, 67 cases), we found D-dimer kept increasing in D-severe group but dropped in the R-severe group after 10 days from the admission onset, and the levels of D-Dimer in these two subgroups were significantly different (14.33 vs. 1.65, *P*<0.001) after 15 days from the admission onset. These results suggested D-dimer might be more related with the CFR compared with other markers. In addition, we checked the case reports of COVID-19 patients and found that D-dimer was also related with the status of patients, and preventive and adjuvant antithrombotic treatment in some severe patients achieved good therapeutic effect (the representative chest X ray images were shown as **Figure 4)**.

**Figure 3:**
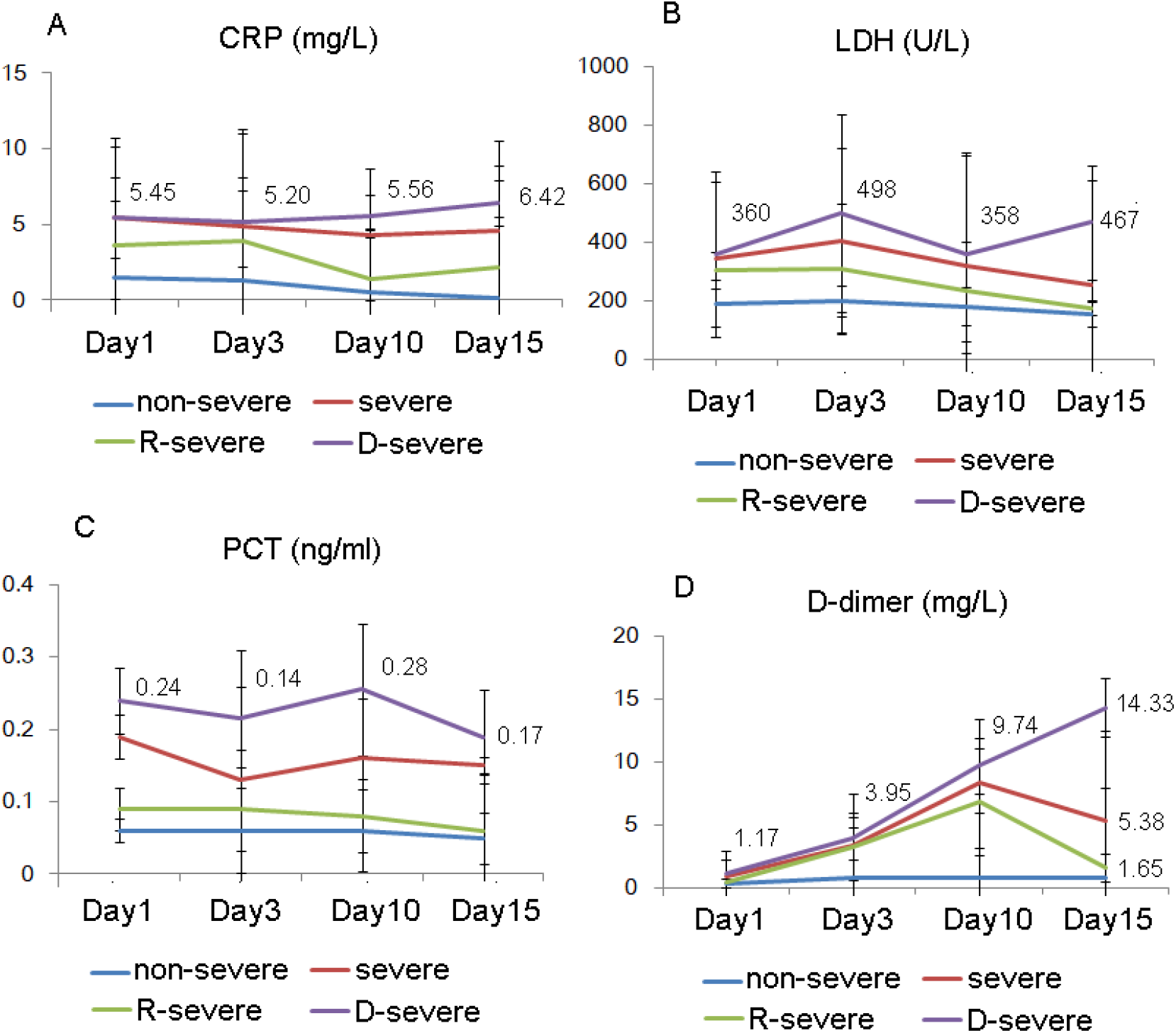
Temporal changes in risk factors between the severe and non-severe patients with COVID-19. Figure shows temporal changes of CRP (A), LDH (B), PCT(C) and D-dimer (D) in patients within 15days **f**rom admission onset. We stratified the patients into 4 subgroups: severe, non-severe R-severe (the severe patients recovered) and D-severe (the patient died). Data represent mean ± SEM. Abbreviations: CRP, C-reactive protein; LDH, lactate dehydrogenase; PCT, procalcitonin.

**Figure 4:**
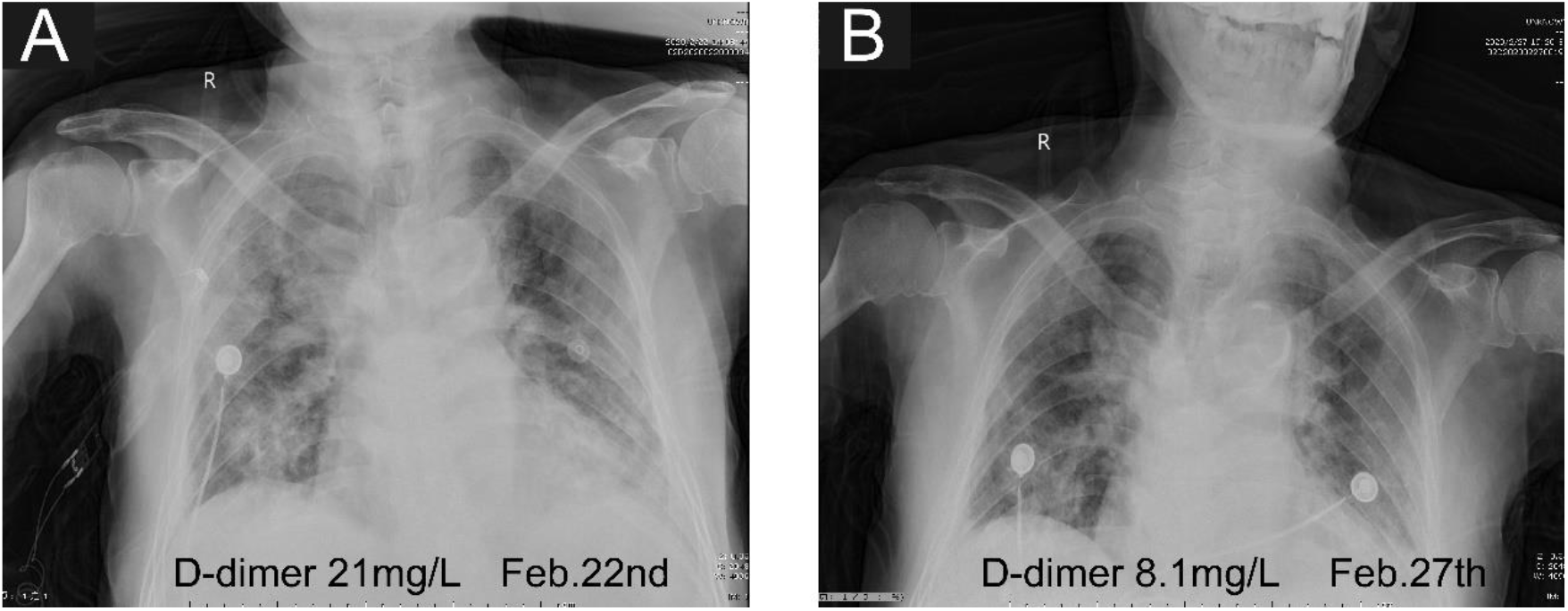
Representative chest X ray images of an 85-year-old male patient during the treatment. A. The chest X-ray suggested that diffuse large area infection of both lungs accompanied, D-dimer (D-D) reached 21.0 mg/l. B. After antiviral therapy and adjuvant antithrombotic treatment with heparin, the bilateral lung infection foci were significantly reduced compared with before, and the level of D-D was found to have subsequently declined to 8.1 mg/l.

## Discussion

Recently, a number of the epidemiological and clinical features of COVID-19 have been reported. Phylogenetic studies indicated that it is closely related to a bat-derived SARS-like coronaviruses (2), and belongs to a large Coronavirus family including severe acute respiratory syndrome coronavirus (SARS)(11) and Middle East Respiratory Syndrome (MERS)(12), which often cause the clinical symptoms ranging from the common fever to severe pneumonia(3). It is reported that COVID-19 has higher infection rates compared with SARS and MERS(13). The median daily reproduction number (R_t_) of COVID-19 in Wuhan declined from 2.35 (95% CI, 1.15-4.77)(13), and the CFR in the severe patient is very high. However, our knowledge about the difference between severe and non-severe COVID-19 patients has not been well established, and the information on prognosis of COVID-19 severity is still lacking. The aim of this study is to elaborate the different Clinical features between severe and non-severe COVID-19 patients, and find the risk factors for predicting the severity of COVIP-19.

In our cohort, we have stratified patients based on the severity on admission according to international guidelines (14). Our study has shown COVID-19 are more severe in elder people, male and the patients with comorbidities. Fever and cough were the dominant symptoms whereas gastrointestinal symptoms were rare. For the diagnosis markers, we found the D-dimer, CRP, LDH, PCT were increased quickly in severe group compared with the non-severe group (CRP, 65.1% vs. 13.5%, *P*<0.001; D-Dimer: 87.3% vs. 35.3%, *P*<0.001; LDH: 83.9% vs. 22.2%, *P*<0.001; PCT: 35.1% vs. 2.2%, *P*<0.001), while ASP, ALP, CK and creatinine were mildly increased. This means more inflammation, thrombus, myocardial injury or liver and kidney damage existed in severe patients. In addition, we made a meta-analysis of 1646 cases with additional 4 related studies to further confirm the risk factors. Moreover, we studied the dynamic changes during the process of COVID-19 by timeline tracking. We found CRP, D-dimer, LDH, PCT kept in higher levels in severe patients. Notably, D-dimer still increased quickly in some severe patients and most likely related with the CFR than other markers. It is noticed that another recent cohort study also showed that D-dimer is a critical risk factors for the CFR of COVID-19 (15).We checked the case reports of severe patients in our cohort retrospectively, and found that prophylactic antithrombotic therapy effectively improved the prognosis in some patients. Thus, we propose that severe patients of COVID-19 can be given preventive anticoagulants or physical therapy. Indeed, when there is serious hypoxemia and hypotension, the possibility of embolism should be considered, and thrombolysis should be timely in the absence of thrombolysis contraindication.

Our study still has several limitations. Firstly, it is reported that some patients with positive chest CT findings may have negative RT-PCR results. In that case, there may be selection bias since only the positive cases confirmed by the RT-PCR were included in our study. Secondly, although we want to track the timeline of each patient’s blood test and other clinical tests, but we did not collect all the data of some patients at each time point for various reasons. This may result in a large standard error of the time axis.

In summary, our study showed CRP, D-dimer, LDH, PCT are the high risk factors related with COVID-19. Especially, D-dimer is a critical indicator for both severity and CFR of COVID-19. As high CFR in the severe COVID-19 patients need to be resolved immediately, the risk factors explored in our study are important of taking into account the disease severity in practice. Though the prognostic factors in our study still need to be further validated by future studies, they should be helpful to provide warning model for predicting severity and mortality in COVID-19, which is very useful for clinical application.

## Methods

### Data sources

We performed a retrospective study on 223 laboratory-confirmed cases with COVID-19 based on clinical indications from 2 hospitals in the Hubei provinces, China, from January 1 through March 8, 2020, were included Wuhan TongJi Hospital and Central Hospital of Wuhan. Initially, suspected cases were diagnosed as having fever, respiratory symptoms or pneumonia. Confirmed case with COVID-19 was defined as a positive result to real-time reverse-transcriptase polymerase-chain-reaction (RT-PCR) assay using nasal and pharyngeal swab specimens(5). The clinical characteristics and laboratory assessments were extracted from electronic medical records. Laboratory assessments consisted of complete blood count, blood chemistry, coagulation test, liver and renal function, C-reactive protein, lactate dehydrogenase and creatine kinase. Radiologic assessments included chest X-ray and computed tomography (CT). The severity of COVID-19 was defined based on the international guidelines for community-acquired pneumonia(14). The ethics committee of Wuhan Tongji Hospital and Central Hospital of Wuhan appoved this study and granted a waiver of informed consent in light of the urgent need to collect clinical data.

### Statistics of the cohort study

Continuous variables were expressed as the means and standard deviations or medians and interquartile ranges (IQR) as appropriate. Categorical variables were summarized as the counts and percentages in each category. We divided COVID-19 patients into severe and non-severe subgroups according to the American Thoracic Society guideline on admission(14). Mann-Whitney test and two-tailed t-test and were applied to continuous variables, the frequencies of categorical variables were compared using the chi-square test and Fisher’s exact test as appropriate. The candidate risk factors included age, sex, abnormal laboratory findings. The tests with p value less than 0.05 was considered statistically significant. Statistical analyses were done using the Statistical Package for the Social Sciences (SPSS) software (version 21.0).

### Meta-analysis

In meta-analysis, we first carried out a computerized literature search of the PubMed, Web of Science, EBSCO and CNKI database (prior to March 15th, 2020) using the following words and terms: ‘‘COVID-19’’, ‘‘SARS-COV-2’’. References of the retrieved publications were also screened. We only selected the literatures which distinguished the clinical characteristics between non-severe and severe COVID-19 patients available with the specific numbers of cases. Then we combined our own cohort study with the data extracted from the selected literatures, and calculated the odd ratio (OR) to assess the association between different risk factors and COVID-19 (**Table 2)**. Chi-square-based *Q*-statistic test was performed to evaluate the between-study heterogeneity of the studies. If P < 0.10, the between-study heterogeneity was considered to be significant, we chose the random-effects model (DerSimonian and Laird model) to calculate the OR, otherwise we chose the fixed-effects model (Mantel–Haenszel model). A pooled OR obtained by meta-analysis was used to give a more reasonable evaluation of the association. A Z test was performed to determine the significance of the pooled OR (P ≤0.05 suggests a significant OR). Analyses were performed by Stata10.0 software.

## Data Availability

The raw/processed data required to reproduce these findings cannot be shared at this time as the data also forms part of an ongoing study.

## Author Contributions

Drs Wu and Wei had full access to all of the data in the study and take responsibility for the integrity of the data and the accuracy of the data analysis. Drs Huang, Y., Lyu, X., and Li, D. contributed equally and share first authorship. Study concept and design: Wu, X., Wei, X., Huang, Y. Acquisition, analysis, or interpretation of data: Wu, X., Huang, Y., Wei, X., Lyu, X., Li, D., Wang, Y., Zou, W. Drafting of the manuscript: Huang, Y., Wu, X., Wei, X., Wang, L., Critical revision of the manuscript for important intellectual content: Huang, Y., Lyu, X., and Li, D. Statistical analysis: Huang, Y., Li, D. Administrative, technical, or material support: Wu, X., Wei, X., Huang, Y., Lyu, X., and Li, D. Study supervision: Wu, X., Wei, X.

## Acknowledgement

We thank Qi Wang, MD (Tongji Hospital), Xin Long, MD (Tongji Hospital) for collecting the data, and Zhongshi Zhou,MD (Hubei University of Chinese Medicine) for preparing the Table. None of these individuals received compensation for their contributions.

